# Liver abnormalities following SARS-CoV-2 infection in children under 10 years of age

**DOI:** 10.1101/2023.09.21.23295905

**Authors:** Pauline Terebuh, Veronica R. Olaker, Ellen K. Kendall, David C. Kaelber, Rong Xu, Pamela B. Davis

## Abstract

**Objective:** Beginning in October 2021 in the US and elsewhere, cases of severe pediatric hepatitis of unknown etiology were identified in young children. While the adenovirus and adenovirus-associated virus have emerged as leading etiologic suspects, we attempted to investigate a potential role for SARS-CoV-2 in the development of subsequent liver abnormalities.

**Design:** We conducted a study utilizing retrospective cohorts of de-identified, aggregated data from the electronic health records of over 100 million patients contributed by US health care organizations.

**Results:** Compared to propensity-score-matched children with other respiratory infections, children aged 1-10 years with COVID-19 had a higher risk of elevated transaminases (Hazard ratio (HR) (95% Confidence interval (CI)) 2.16 (1.74-2.69)) or total bilirubin (HR (CI) 3.02 (1.91-4.78)), or new diagnoses of liver diseases (HR (CI) 1.67 (1.21-2.30)) from one to six months after infection. Patients with pre-existing liver abnormalities, liver abnormalities surrounding acute infection, younger age (1-4 years), or illness requiring hospitalization all had similarly elevated risk. Children who developed liver abnormalities following COVID-19 had more pre-existing conditions than those who developed abnormalities following other infections.

**Conclusion:** These results indicate that SARS-CoV-2 may prime the patient for subsequent development of liver infections or non-infectious liver diseases. While rare (∼1 in 1,000), SARS-CoV-2 is a risk for subsequent abnormalities in liver function or the diagnosis of diseases of the liver.

**What is already known on this topic:** Clusters of severe hepatitis in children in 2022 coincident with the increase in COVID-19 infections in children raised the question of the contribution of SARS-CoV-2 to the hepatitis outbreak, though it was soon determined that SARS-CoV-2 was not the primary etiologic agent.

**What this study adds:** SARS-CoV-2 may prime the patient for subsequent development of liver infections or non-infectious liver diseases.

**How this study might affect research, practice or policy:** Despite the mild initial disease in children, there may be longer term consequences of COVID-19, such as liver abnormalities, that warrants further investigation.

## INTRODUCTION

In early 2022, reports emerged of clusters of severe hepatitis not associated with any of the usual viruses in young children in Scotland and soon after, the United States and elsewhere in the United Kingdom. [1-5] The coincidence in timing of this outbreak with the rise of childhood cases of COVID-19 as the Omicron variant spread made SARS-CoV-2 a tempting explanation for this new disease. For other viral infections, such as influenza, EBV, Coxsackie B, and SARS/MERS, damage to the liver is well-documented, and is often considered to result from T-cell mediated attack without necessarily the presence of the virus or viral antigens in the liver. [6] In addition, many viruses are known to leave patients vulnerable to secondary invaders. However, it soon became apparent that, although SARS-CoV-2 antibodies could be demonstrated in over half the patients, it was not possible to implicate the virus directly in severe liver damage. [7] Subsequent studies have suggested that the culprit in the outbreak of severe hepatitis might be adenovirus type 41 subtype F (Ad41F) or Ad 41F plus adeno-associated virus 2 (AAV2). [8,9] Some series indicate that as many as 80% of patients have evidence of infection with Ad41, [2] so despite the absence of the usual pathologic signature of adenoviral hepatitis, inclusion bodies, this explanation seems the most likely. The reason for the sudden outbreak at widely separated geographic locations, however, is unclear. Concern persists that our lack of understanding of this outbreak may leave us less able to cope effectively with another such episode, should it occur.

The timing of the hepatitis outbreak coincident with the surge in COVID-19 in children suggested some role for SARS-CoV-2 in priming children for the devastating outbreak. [10] In adults, SARS-CoV-2 is known to produce lingering alterations in the immune system that can also lead to autoimmune phenomena. [11, 12] To investigate the possible role of SARS-CoV-2 in liver dysfunction in children, we examined the risk for protracted abnormalities in liver enzymes or bilirubin, or new diagnoses of hepatitis or liver disease, in children following diagnosis of COVID-19 compared to children who had other respiratory infections.

## METHODS

We conducted a study utilizing retrospective cohorts of aggregated and deidentified electronic health record (EHR) data from over 100 million patients on the TriNetX analytics platform contributed by 59 health care organizations (HCOs) in the United States (US) representing a diverse population geographically, demographically and in insurance status. We employed the US Collaborative Network for the primary analyses. For additional privacy protection, some HCOs that contribute to the network date-shift individual EHRs from 1-365 days on the calendar while maintaining the relative timing of clinical data within each EHR. Analyses investigating secular trends during the pandemic or comparing periods predominated by SARS-CoV-2 variants (Delta, Omicron) were performed in the Research USA Minimal Date Shift Network including only HCOs that contributed minimally shifted EHR data (0-7days). Built-in analytic functions allow for analysis of patient-level data in aggregate form only, without individual protected health information. See Supplement for details. The MetroHealth System Institutional Review Board has designated studies using TriNetX in ways such as described in this manuscript, as exempt. The study design and results reporting followed the guidelines of the Strengthening the Reporting of Observational Studies in Epidemiology (STROBE).

### Cohorts

The study population was comprised of children aged 1 to 10 years at the time of the diagnosis of a respiratory infection (COVID-19 or a non-COVID-19 respiratory infection) between January 1, 2020 and December 31, 2022. The lower age limit was selected to avoid perinatal complications and upper age limit was selected because reports of the surge in hepatitis of unknown origin in children reported very few cases above the age of 10 years. A sub-population of children aged 1 to 4 years was also analyzed because most cases described during the outbreak were in this age range. Criteria for inclusion in the COVID-19 cohort had either *International Statistical Classification of Diseases and Related Health Problems, Tenth Revision* (ICD-10) code for COVID-19 (U07.1), J12.81 (pneumonia due to SARS-associated coronavirus), or J12.82 (pneumonia due to coronavirus disease 2019) as an encounter diagnosis, or a positive laboratory result for SARS-CoV-2 RNA detection (see Supplement for codes included the composite code). Inclusion in the non-COVID-19 other respiratory infection (ORI) cohort required an encounter diagnosis of J00-J06 (other acute upper respiratory infections), J20-J22 (other acute lower respiratory infections), or J09-J18 (influenza and pneumonia) and no record of COVID-19, positive SARS-CoV-2 RNA detection nor SARS-CoV-2 antibodies during the pre-vaccine period (see Supplement for details). Patients were excluded from both cohorts for pre-existing liver disease documented by an ICD-10 encounter diagnosis of E88.01 (alpha-1-antitrypsin deficiency), B15-B19 (viral hepatitis), K70-K77 (diseases of the liver), or elevated serum laboratory values (aspartate aminotransferase (AST) ≥110 units per liter (U/L), alanine aminotransferase (ALT) ≥100 U/L, or total bilirubin (TBil) ≥2 milligrams/deciliter (mg/dl)) up until one month prior to the first record of the respiratory infection encounter diagnosis. To further investigate whether COVID-19 impacts patients with pre-existing liver disease, additional cohorts were constructed without excluding patients with pre-existing conditions. See Supplement for details.

To further examine the subset of patients in the cohorts with disease severity sufficient to require hospitalization, sub-cohorts were constructed including only patients with a documented inpatient stay during the acute period (from 1 month prior to 1 month after) the COVID-19 or ORI diagnosis. Additionally, the subset of patients who had AST/ALT or TBili measured during the acute period were compared to controls for access to laboratory testing.

Using the Research USA Minimal Date Shift network, five serial 6-month cohorts were constructed: April 1, 2020 to September 30, 2020; October 1, 2020 to March 31, 2021; April 1, 2021 to September 30, 2021; October 1, 2021 to March 31, 2022; April 1, 2022 to September 30, 2022 to evaluate secular trends. Additional cohorts corresponding to patients with infections that occurred during variant waves were constructed to index a patient’s first documented COVID-19 infection or corresponding non-COVID-19 infection occurring when virologic surveillance data showed a prevalence of >90% for the respective virus variant: Delta, August 1, 2021 to November 30, 2021; Omicron, December 26, 2021 to September 30, 2022.

To describe and compare the baseline characteristics of patients who developed liver complications during the 1–6-month follow-up period, additional inclusion criteria were added to existing cohort definitions: ICD-10 codes for the outcomes under study (K70-K77 or B15-B19) occurring during the 1–6-month follow-up period. See below for more details on these outcome diagnostic codes.

### Outcomes

Evidence of liver disease was identified by ICD-10 encounter diagnoses: K70-K77 (diseases of the liver), B15-B19 (viral hepatitis), or elevation in serum laboratory values reflecting transaminitis (AST ≥110 U/L or ALT ≥100 U/L) or cholestasis (TBil ≥2 mg/dl). These outcomes were intended to be comprehensive to capture all acute hepatic complications. Subsets of these outcomes were further analyzed by grouping specific ICD-10 codes. The National Syndromic Surveillance Program and the Premier Healthcare Database Special Release used the following codes to identify hepatitis of unspecified etiology [13]: B17.8 (other specified acute viral hepatitis); B17.9 (acute viral hepatitis, unspecified); B19.0 (unspecified viral hepatitis with hepatic coma); B19.9 (unspecified viral hepatitis without hepatic coma); K71.6 (toxic liver for liver disease with hepatitis, not elsewhere classified); K72.0 (acute or subacute hepatic failure); K75.2 (nonspecific reactive hepatitis). We used those codes as a composite outcome with one additional code used for outbreak case finding: K75.9 (inflammatory liver disease, unspecified). [2] Other composite outcome subsets included K70-K77 (diseases of the liver), B15-B19 (viral hepatitis), and specific codes for acute hepatic complications for which counts were sufficient for analysis: K72 (hepatic failure, not elsewhere classified); K75 (other inflammatory liver diseases); K76 (other diseases of liver).

### Statistical analysis

Statistical analyses were performed during August to September 2023, using the TriNetX analytics platform. We compared outcomes between cohorts after propensity-score matching (1:1 matching using a nearest neighbor greedy algorithm with a caliper of 0.25 times the standard deviation) for demographic factors including age, sex, race, and ethnicity; pediatric body mass index (BMI); encounter for preventive medicine services (based on CPT codes); encounter for immunizations (based on CPT codes); and SARS-CoV-2 vaccination as recorded one day prior to the first encounter for the respiratory illness. See supplement for specific codes used for matching. A standard mean difference (SMD) of ≤0.1 was considered a good match. Reports of COVID-19 vaccination status are likely incomplete as many patients received vaccination in other settings, which may not be recorded in the EHR.

We compared outcomes between matched cohorts during the 1-to-6-month period following the index event (COVID-19 or ORI) after excluding patients with the outcome during the acute period (from 1 month prior to 1 month after the encounter diagnosis of COVID-19 or ORI). Built-in TriNetX analytics use the Cox proportional hazard model to calculate hazard ratios (HR) and 95% confidence intervals (CI). The proportional hazard assumption is tested using the generalized Schoenfeld approach. To protect patient privacy when the aggregate counts for a reported outcome are small, i.e., ≤10, the TriNetX platform reports the count as 10. However, in generating the Kaplan-Meier curves and calculating hazard ratios, the TriNetX platform uses actual counts in calculating the rates even when the count is less than 10. Patients are censored from the survival analysis after the last clinical fact in the EHR. Since patients in the study cohorts who had outcomes recorded during both the acute period and also during the 1–6-month follow-up period were excluded in the primary analysis, an additional comparison was made without excluding those patients to determine whether the elevated liver enzymes or bilirubin values could have been persistent from the acute phase. Relative risks (RR) and 95% confidence intervals (CI) for the outcomes during the 1–6-month follow-up period were calculated (Kaplan-Meier survival analysis and HR was not used because the outcome may have already occurred during the acute period). Small counts ≤10 are reported as 10. The RR calculation uses 10 (in contrast to the Kaplan-Meier survival analysis that uses actual counts), so may underestimate RR when one of the cohorts has a count of less than 10. Additional analyses comparing cohorts without excluding those with pre-existing liver abnormalities employed RR as well because the outcome may have occurred previously.

## RESULTS

The overall study population was comprised of children 1-10 years of age without pre-existing liver disease with EHR-documented COVID-19 (263,069) or a non-COVID-19 respiratory infection (1,111,362) between January 1, 2020 to December 31, 2022. The study population was drawn from 5,660,153 similarly aged children with at least one encounter during the study period. Prior to matching, patients in the COVID-19 cohort were significantly older than patients in the ORI cohort (Table 1). After propensity matching on baseline characteristics (Table 1), 260,132 patients were in each cohort. Tables summarizing baseline characteristics for the subset of children aged 1-4 years, and the subsets of children in both age groups with a record of hospitalization during the acute period (from 1 month prior to 1 month after the respiratory infection encounter diagnosis) can be found in the Supplement.

**Table 1.** Baseline Characteristics of Study Population Before and After Matching (Age 1-10 Years)

|  | Unmatched cohort, No. (%) |  |  | Matched cohort, No. (%) |  |  |
| --- | --- | --- | --- | --- | --- | --- |
|  | COVID-19<br>n=263,069 | ORI<br>n=1,111,362 | SMD | COVID-19<br>n=260,132 | ORI<br>n=260,132 | SMD |
| Age at Index in years, mean (SD) | 5.17±3.00 | 4.46±2.87 | 0.24 | 5.17±3.00 | 5.16±3.00 | 0.003 |
| Sex |  |  |  |  |  |  |
| Male | 137,041 (52.7) | 572,243 (52.1) | 0.01 | 137,041 (52.7) | 136,860 (52.6) | 0.001 |
| Female | 122,928 (47.3) | 525,718 (47.9) | 0.01 | 122,928 (47.3) | 123,177 (47.4) | 0.002 |
| Unknown | 165 (0.1) | 346 (<0.1) | 0.01 | 163 (0.1) | 95 (<0.1) | 0.012 |
| Ethnicity |  |  |  |  |  |  |
| Not Hispanic | 159,371 (61.3) | 677,379 (61.7) | 0.01 | 159,369 (61.3) | 159,841 (61.4) | 0.004 |
| Hispanic | 43,986 (16.9) | 187,765 (17.1) | <0.01 | 43,986 (16.9) | 44,086 (16.9) | 0.001 |
| Unknown | 56,777 (21.8) | 233,163 (21.2) | 0.01 | 56,777 (21.8) | 56,225 (21.6) | 0.005 |
| Race |  |  |  |  |  |  |
| White | 138,595 (53.3) | 598,920 (54.5) | 0.03 | 138,595 (53.3) | 139,075 (53.5) | 0.004 |
| Black | 52,889 (20.3) | 198,847 (18.1) | 0.06 | 52,887 (20.3) | 52,317 (20.1) | 0.006 |
| Asian | 8989 (3.5) | 40,190 (3.7) | 0.01 | 8989 (3.5) | 8949 (3.4) | 0.001 |
| Native American or Alaska Native | 1229 (0.5) | 6154 (0.6) | 0.01 | 1229 (0.5) | 1166 (0.5) | 0.004 |
| Native Hawaiian or Other Pacific Islander | 526 (0.2) | 2677 (0.2) | 0.01 | 526 (0.2) | 477 (0.2) | 0.004 |
| Unknown Race | 57,906 (22.3) | 251,519 (22.9) | 0.02 | 57,906 (22.3) | 58,148 (22.4) | 0.002 |
| BMI, pediatric |  |  |  |  |  |  |
| measured | 36,455 (14.0) | 165,475 (15.1) | 0.03 | 36,455 (14.0) | 36,692 (14.1) | 0.003 |
| <5<sup>th</sup> percentile | 2753 (1.1) | 11,853 (1.1) | <0.01 | 2735 (1.1) | 2576 (1.0) | 0.006 |
| 5<sup>th</sup> to <85<sup>th</sup> percentile | 24,003 (9.2) | 116,910 (10.6) | 0.05 | 24,003 (9.2) | 24,124 (9.3) | 0.002 |
| 85<sup>th</sup> to <95<sup>th</sup> percentile | 7742 (3.0) | 32,619 (3.0) | <0.01 | 7742 (3.0) | 7714 (3.0) | 0.001 |
| ≥95<sup>th</sup> percentile | 10,623 (4.1) | 38,525 (3.5) | 0.03 | 10,623 (4.1) | 10,660 (4.1) | 0.001 |
| Preventive medicine services | 99,104 (38.1) | 450,554 (41.0) | 0.06 | 99,104 (38.1) | 99,720 (38.3) | 0.005 |
| Encounter for immunization | 88,630 (34.1) | 417,136 (38.0) | 0.08 | 88,630 (34.1) | 88,873 (34.2) | 0.002 |
| SARS-CoV-2 vaccination | 2365 (0.9) | 6463 (0.6) | 0.04 | 2365 (0.9) | 2065 (0.8) | 0.013 |
| Abbreviations: ORI, other respiratory infection; SMD, standard mean difference; SD, standard deviation |  |  |  |  |  |  |

Table 2 summarizes the counts and hazard of hepatic complications after COVID-19 vs ORI for both age groups (1-4 years, 1-10 years) for all cases and for the subset with an associated hospitalization. During the 1-6 months after the encounter diagnosis for the respiratory infection, patients with COVID-19 had a significantly greater hazard of elevated AST or ALT, and TBili with HR point estimates ranging from 2.13-4.40. HR point estimates were significantly increased for the composite outcome of diseases of the liver or viral hepatitis (K70-K77, B15-19: HR point estimate range, 1.48-4.50), and for diseases of the liver alone (K70-K77: HR point estimate range, 1.67-4.38). Few encounters were coded with the following ICD-10 encounter diagnoses: alcoholic liver disease (K70); toxic liver disease (K71); hepatic failure, not elsewhere classified (K72) (except for all patients aged 1-10 years with 12 in the COVID-19 cohort and 10 in the ORI cohort (HR (CI) 11.42 (1.49-87.83))); chronic hepatitis, not elsewhere classified (K73); fibrosis and cirrhosis of liver (K74). ICD-10 terms with sufficient counts to allow comparison had elevated HRs: other inflammatory diseases of the liver (K75: HR point estimate range, 2.85-9.85) which includes autoimmune hepatitis (K75.4) and other specified inflammatory liver diseases (K75.8); and other diseases of the liver (K76: HR point estimate range, 1.46-3.97) which includes fatty change of liver, not elsewhere classified (K76.0). The composite outcome of ICD-10 codes used for surveillance of hepatitis of unspecified etiology surveillance also had significantly elevated HRs (HR point estimate range, 2.62-6.23). All HRs had 95% CIs that were greater than 1 except the outcome of other inflammatory diseases of the liver in the 1–4-year age group. The Cox proportionality assumption at p≥0.05 was met for all comparisons except elevated ALT/AST for 1–4-year age group with inpatient stay (p=0.049), and elevated TBil for 1–10-year-old age group with inpatient stay (p=0.01). A comparison of these outcomes without excluding patients with the encounter diagnosis for one these outcomes during the acute period had increased RR estimates for all comparisons. See Supplement eTable 4. Analyses conducted with cohorts of patients without excluding patients with pre-existing liver abnormalities showed similar results. Furthermore, additional ICD-10 terms had sufficient counts to report from this analysis, showing an elevated risk in the COVID-19 cohort for viral hepatitis (B15-B19), and hepatic failure, not otherwise classified (K72). See Supplement eTable 5.

**Table 2.**
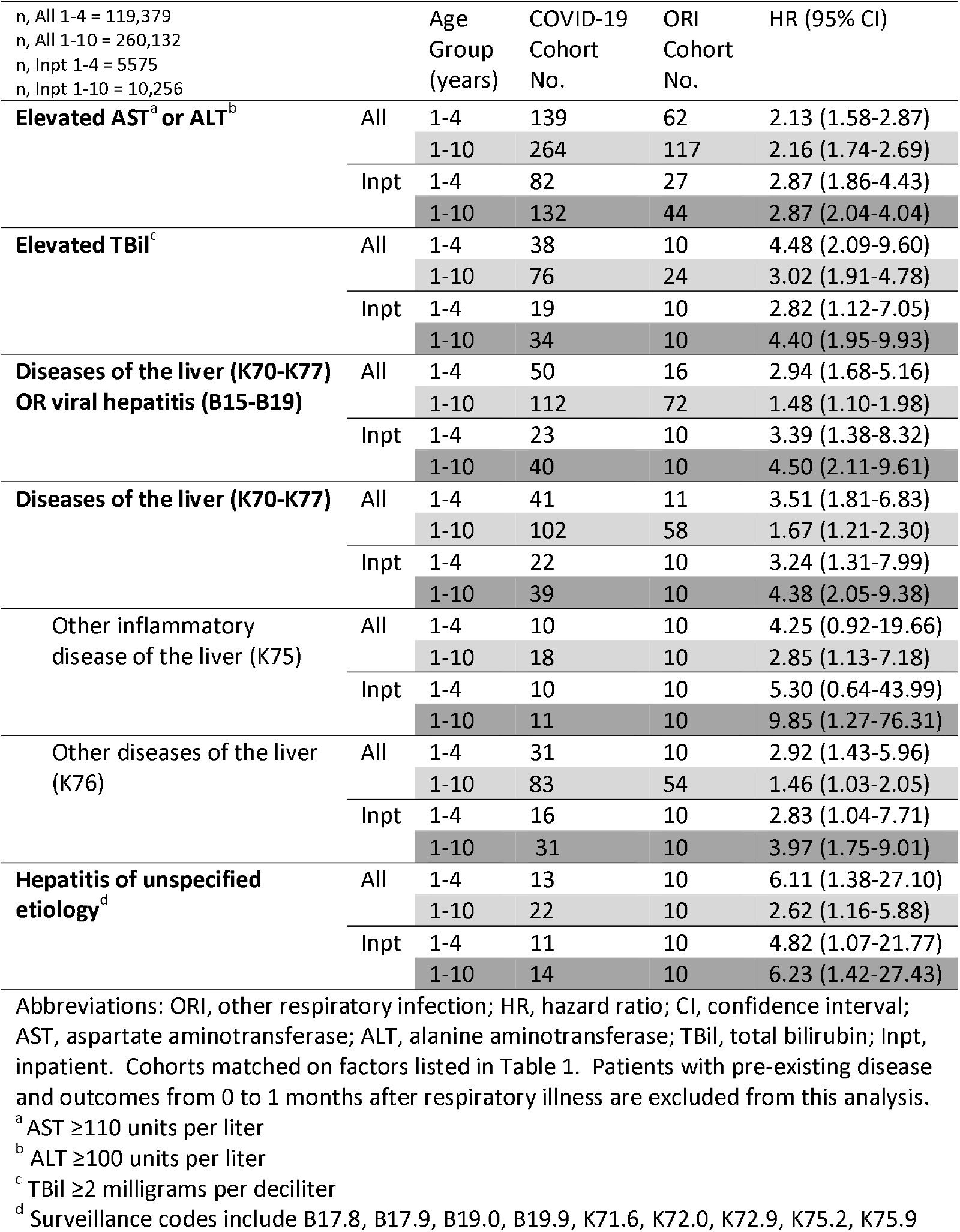
Comparison of clinical outcomes during 1 to 6 months after COVID-19 vs ORI among matched cohorts of patients aged 1-10 years (01/01/2020 – 12/31/2022)

When comparison was restricted to patients with AST, ALT or TBil measured, but not shown to be elevated during the acute period, patients with COVID-19 had a greater hazard for elevated AST, ALT, or TBil during the 1-6-month follow-up period in both age groups (Table 3). Similarly, a comparison was made between patients with labs measured during the acute period, but without excluding those who may have had elevated labs during the acute period. The RR for persistent or newly incident lab elevations was greater for patients after COVID-19 (eTable 6). Among patients aged 1-10 years with persistent lab elevations into the 1–6-month follow-up period after COVID-19, the most recent elevated lab value mean U/L (SD) were as follows: AST 236 (1760); ALT 133 (260); TBil 2.16 (2.89).

**Table 3.**
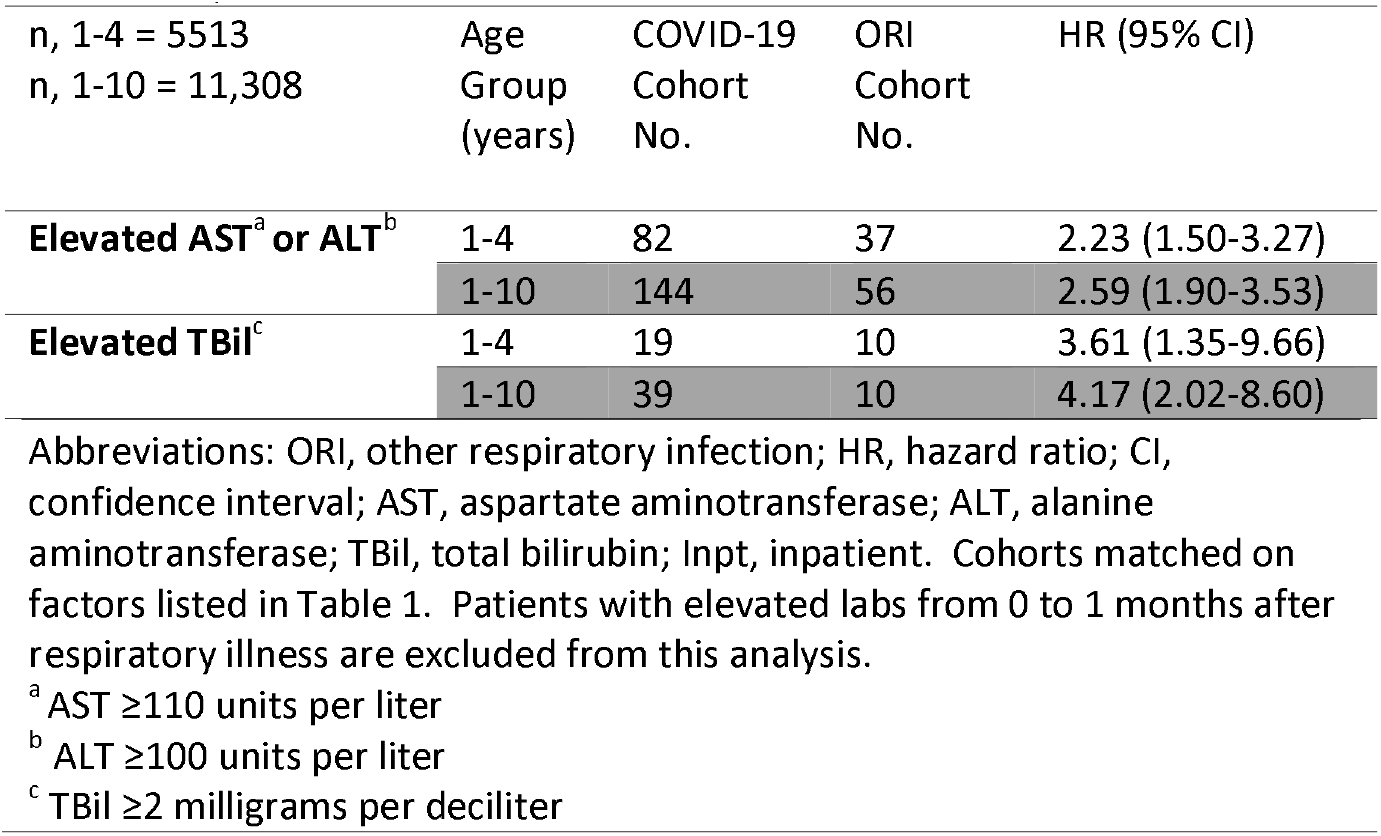
Comparison of elevated laboratory values during 1 to 6 months after COVID-19 vs ORI among matched cohorts of pediatric patients with laboratory values measured during the acute period of infection (01/01/2020 – 12/31/2022)

| n, 1-4 = 5513<br>n, 1-10 = 11,308 | Age<br>Group<br>(years) | COVID-19<br>Cohort<br>No. | ORI<br>Cohort<br>No. | HR (95% CI) |
| --- | --- | --- | --- | --- |
| **Elevated AST<sup>a</sup> or ALT<sup>b</sup>** | 1-4 | 82 | 37 | 2.23 (1.50-3.27) |
|  | 1-10 | 144 | 56 | 2.59 (1.90-3.53) |
| **Elevated TBil<sup>c</sup>** | 1-4 | 19 | 10 | 3.61 (1.35-9.66) |
|  | 1-10 | 39 | 10 | 4.17 (2.02-8.60) |

Abbreviations: ORI, other respiratory infection; HR, hazard ratio; CI, confidence interval; AST, aspartate aminotransferase; ALT, alanine aminotransferase; TBil, total bilirubin; Inpt, inpatient. Cohorts matched on factors listed in Table 1. Patients with elevated labs from 0 to 1 months after respiratory illness are excluded from this analysis.
<sup>a</sup> AST ≥110 units per liter
<sup>b</sup> ALT ≥100 units per liter
<sup>c</sup> TBil ≥2 milligrams per deciliter

The prevalence of pre-existing conditions (documented at least a month before the respiratory diagnosis) among patients who, during the 1–6-month follow-up period, had encounter diagnoses for diseases of the liver (K7-K77) or viral hepatitis (B15-B19) following COVID vs ORI were compared. Cases with hepatic complications that followed COVID-19 vs ORI were older (mean, 5.51 vs 4.42 years) and had a higher prevalence of pre-existing conditions. However, when limiting the comparison to cases who were hospitalized during the acute phase of the respiratory infection, the prevalence of pre-existing conditions among COVID-19 or ORI associated cases varied by condition. See eTable 7 for details.

Analyses comparing shorter time periods yielded smaller total counts and did not uncover any temporal or variant-related trends. These analyses were conducted using the Research USA Minimally Shifted Network which has approximately 35% fewer records. When comparing COVID-19 to ORI during 5 serial 6-month time blocks from 03/01/2020 to 09/30/2022, there were no significant secular trends. Elevations in liver enzymes and bilirubin were evident in all time periods. Liver-related complications were seen in each time period, however, the relatively small counts when divided over 5 time periods made the comparison between COVID-19 and ORI underpowered. When restricting analyses to infections that occurred during COVID-19 waves in which either the Delta or the Omicron variant was predominant, results for liver enzymes and bilirubin were similar for the COVID-19 to ORI cohort comparison during each wave as when comparing the results over the entire study period. No differences in liver enzyme and bilirubin elevation were found when comparing COVID-19 cohorts between waves.

## DISCUSSION

In a large population sample from across the US, children who had COVID-19 were at significantly higher risk for elevated liver enzymes and bilirubin or a new diagnosis of hepatitis of various types in the following six months. These elevations were observed for children aged 1 to 10 years and also for younger children aged 1 to 4 years when compared to patients with a non-COVID-19 respiratory infection, and it was true for patients with COVID-19 of unspecified severity, as well as for those with an associated hospitalization. The risk was increased during the period 1 to 6 months after COVID-19 diagnosis whether or not patients with pre-existing liver disease were excluded or whether or not patients with liver abnormalities during the acute illness period were excluded from the sample. The risk was also noted when comparing only patients with normal labs measured during the acute phase of the illness. The increased risk for elevated transaminases or bilirubin was evident following a COVID-19 diagnosis during each time period of the pandemic. Because the surge in severe hepatitis among pediatric patients occurred coincident with the Omicron variant surge in the pandemic, we directly compared the time when the Delta variant predominated to the time when over 90% of sequenced viruses were the Omicron variant. No difference in the risk of subsequent liver enzyme or bilirubin elevations were found between variant waves.

Since several respiratory infections are known to be associated with subsequent liver abnormalities, the excess of new cases among children who contracted SARS-CoV-2 compared to those with prior respiratory infections may be especially telling. Indeed, the excess risk of elevated transaminases or bilirubin was much higher when children diagnosed with COVID-19 were compared to children who had a medical encounter for any reason, but this category could include well child visits, fractures, or allergies, diagnoses much less likely to display liver abnormalities than children who had viral respiratory infections. However, even compared to other respiratory infections, COVID-19 posed more of a risk for subsequent liver diagnoses, suggesting that it does impose particular vulnerability.

We tested whether prior infection with SARS-CoV-2 is associated with subsequent diagnoses of hepatitis of known viral origin (hepatitis A-E), unspecified viral hepatitis, or cases of acute liver complications for which an infectious origin was not diagnosed, including toxic liver disease, hepatic failure, other inflammatory diseases of the liver (including autoimmune hepatitis), or other diseases of the liver (including fatty change). We reasoned that if COVID-19 conferred vulnerability by any of the mechanisms involving the immune system, this vulnerability may extend to many infectious or even non-infectious disorders of the liver. For all acute disease ICD-10 codes examined for which there was a large enough number of cases for comparison, the risk was higher among children 1-6 months following COVID-19 than it was following other respiratory infections. For the following codes: viral hepatitis (B15-B19); toxic liver disease (K71); hepatic failure, not elsewhere classified (K72); the number of cases was too small for the primary analysis. We speculate that ongoing low grade liver inflammation following COVID-19, though rare (approximately 10 in 10,000) among children without a history of liver abnormalities, leaves children vulnerable to either develop liver disease or to acquire an additional infection. Nevertheless, if 10 million children were infected, 10,000 or more might be left at risk.

Children were more likely to have liver enzymes or bilirubin measured during the following 1-6 months if they were diagnosed with COVID-19 at the index encounter (RR, 1.8) even though the risk of a follow-up visit for any reason was only modestly increased for children with COVID-19 (RR, 1.07). This probably speaks to the index of suspicion in the physicians caring for these children. However, this excess of sampling might create some bias of ascertainment in detecting liver abnormalities among children following COVID. Therefore, we compared only those children who had blood drawn to measure liver enzymes or bilirubin around the time of the diagnosis of COVID-19 or other respiratory infection. For this comparison, laboratory measurements showing liver enzymes and bilirubin were also elevated among children following COVID-19.

How COVID-19 might predispose to later liver abnormalities is a matter for speculation. For adults, four mechanisms have been suggested: direct infection, toxicity from cytokine storm or hypoxic injury associated with severe COVID-19, toxicity from the drugs used to treat COVID-19, and immune phenomena. [14] Direct infection of hepatocytes is unlikely because viral receptors are sparse on hepatocytes, although they do occur in abundance on cholangiocytes. However, there are case reports of acute hepatitis associated with COVID-19, some of them accompanying MIS-C, and others, without significant respiratory involvement. [15] Though in adults, worse acute liver injury is associated with more severe COVID-19, we did not observe an association with severity of COVID-19 as assessed by need for hospitalization. Moreover, children tend to have less severe disease than adults. Therefore, it is less likely that cytokine storm, vascular injury, or hypoxia during the acute illness account for our findings. Few antiviral drugs are approved for children under 10 years of age, so drug induced hepatotoxicity is also less likely than in adults. Therefore, immune phenomena seem the most likely explanation. Many autoimmune phenomena have been documented following SARS-CoV-2 infection, so an autoimmune mechanism of liver damage is possible. [16] Case reports of autoimmune hepatitis following COVID-19 in adults have appeared. [17-19] In addition, for other viruses, T-cell mediated damage to the liver has been observed even in the absence of persistence of the virus or viral antigens in the liver. [20] Further work is needed to sort out these possibilities.

### Limitations

This study has several limitations. Its retrospective observational design precludes determination of causality, only association. Especially during the later time periods of the pandemic, it is likely that some patients with COVID-19 were not medically attended nor tested for SARS-CoV-2 in a way recorded in the EHR. Therefore, some patients who had COVID-19 may have been included with the ORI control group. However, such an error would tend to reduce the group differences rather than create them. Not all patients diagnosed with liver diseases had liver enzymes or bilirubin recorded in TriNetX, either because the tests were performed outside of the health care organization, or because at some health care organizations, laboratory databases are not included in the TriNetX uploads. Therefore, an exact match of the biochemical abnormalities and the diagnoses is not possible. In addition, the TriNetX database, though quite large and covering about 28% of the US population, may not be entirely representative. Further studies focusing in on the key issues raised by this report are warranted.

For patients hospitalized around the time of the encounter diagnosis for COVID-19 or non-COVID-19 encounter diagnosis, the indication for hospitalization cannot be ascertained, but likely represents severe COVID-19. Nevertheless, hospitalized patients with these concurrent respiratory infections had a similar prevalence of common underlying conditions, but a substantial difference in the subsequent risk for persistent liver disease as documented by encounter diagnoses or elevation of AST/ALT or TBili.

## Data Availability

All data produced in the present work are contained in the manuscript

## ACKNOWLEDGEMENTS

Conflicts of Interest and Sources of Funding: No author declares a conflict of interest. This study was supported by grants from the National Institutes of Health AG057557, AG061388, AG062272, AG076649 (all to RX), AA029831 and AG076649 to RX and PBD, and grant 1UL1TR002548-01 to Michael Konstan, which provided core infrastructure support.

## Notes

### Competing Interest Statement

The authors have declared no competing interest.

### Author Declarations

The MetroHealth Institutional Review Board has designated this study as exempt.

