## Supplement for "Liver abnormalities following SARS-CoV-2 infection in children under 10 years of age"

**eMethods**

**eTable 1.**

**eTable 2.**

**eTable 3.**

**eTable 4.**

**eTable 5.**

**eTable 6.**

**eMethods**

**TriNetX Database and Statistical Analysis Description of TriNetX database**: The data used in this study were accessed from August to September 2023 from the TriNetX US Collaborative Network without the use of natural language processing or the Research USA Minimal Date Shift Network with date shifting of records limited to 0-7 days. This resource provides access to electronic health records (diagnoses, procedures, medications, laboratory values, genomic information) from over 100 million patients from 59 healthcare organizations, which is de-identified per criteria from the Health Insurance Portability and Accountability Act (HIPAA), Section §164.514(a) of the HIPAA Privacy Rule. MetroHealth System, Cleveland, Ohio, IRB has determined that research using TriNetX in ways such as described in this manuscript, is not Human Subject Research and therefore exempt from IRB review.

The TriNetX platform de-identifies and aggregates electronic health record (EHR) data from 59 contributing healthcare systems, most of which are large academic medical institutions with both inpatient and outpatient facilities at multiple locations, across all 50 states in the US. Patient EHR data includes information from hospitals, primary care, and specialty treatment providers, covering diverse geographic locations, age groups, racial and ethnic groups, income levels and insurance types including various commercial insurances, governmental insurance (Medicare and Medicaid), self-pay/uninsured, worker compensation insurance, and military/VA insurance among others. Race and ethnicity data in TriNetX is derived from self-reports in the clinical EHR systems, which is then mapped to the following categories: (1) Race: Asian, American Indian or Alaskan Native, Black or African American, Native Hawaiian or Other Pacific Islander, White, Unknown race; and (2) Ethnicity: Hispanic or Latino, Not Hispanic or Latino, Unknown Ethnicity.

COVID-19 Cohort Inclusion and Exclusion Criteria Codes

Included if any of code below:

U07.1 COVI-19

J12.81 Pneumonia due to SARS-associated coronavirus

J12.82 Pneumonia due to coronavirus disease 2019

9088 (TNX curated) SARS coronavirus 2 and related RNA [Presence] – positive

Excluded if any code below one or more months before the first above code:

E88.01 Alpha-1-antitrypsin deficiency

B15-B19 Viral hepatitis

K70-K77 Diseases of the liver

9047 (TNX curated) Aspartate aminotransferase {Enzymatic activity/volume] in Serum or Plasma ≥ 110 U/L

9044 (TNX curated) Alanine aminotransferase [Enzymatic activity/volume] in Serum, Plasma or Blood ≥ 100 U/L

9050 (TNX curated) Bilirubin total [Mass/volume] in Serum, Plasma or Blood ≥2mg/dL

ORI Cohort Inclusion and Exclusion Criteria Codes

Included if any of code below:

J00-J06 Acute upper respiratory infections

J09-J18 Influenza and pneumonia

J20-J22 Other acute lower respiratory infections

Excluded if any code below one or more months before the first above code:

E88.01 Alpha-1-antitrypsin deficiency

B15-B19 Viral hepatitis

K70-K77 Diseases of the liver

9047 (TNX curated) Aspartate aminotransferase {Enzymatic activity/volume] in Serum or Plasma ≥ 110 U/L

9044 (TNX curated) Alanine aminotransferase [Enzymatic activity/volume] in Serum, Plasma or Blood ≥ 100 U/L

9050 (TNX curated Bilirubin total [Mass/volume] in Serum, Plasma or Blood ≥2mg/dL

Excluded if any code below occurred between January 20, 2020 and March 31, 2020

B97.29 Other coronavirus as the cause of diseases classified elsewhere

B34.2 Coronavirus infection, unspecified

J12.81 Pneumonia due to SARS-associated coronavirus

Excluded if any code below occurred anytime

J12.82 Pneumonia due to coronavirus disease 2019

U07.1 COVID-19

9088 (TNX curated) SARS coronavirus 2 and related RNA [Presence] - positive

Excluded if any code below occurred on or before October 29, 2021

9089 (TNX curated) SARS coronavirus 2 IgG IgM Ab [Presence] in Serum or Plasma – positive

94505-5 SARS-CoV-2 (COVID-19) IgG Ab [Units/volume] in Serum or Plasma by Immunoassay ≥0.1 [arbU]/ml, ever

94506-3 SARS-CoV-2 (COVID-19) IgM Ab [Units/volume] in Serum or Plasma by Immunoassay ≥0.1 [arbU]/ml, ever

94562-6 SARS-CoV-2 (COVID-19) IgA Ab [Presence] in Serum or Plasma by Immunoassay – positive, ever

94762-2 SARS-CoV-2 (COVID-19) Ab [Presence] in Serum or Plasma by Immunoassay – positive, ever

94769-7 SARS-CoV-2 (COVID-19) Ab [Units/volume] in Serum or Plasma by Immunoassay ≥0.1 [IU]/ml, ever

TNX curated 9088 is a composite code created by TriNetX and includes RNA detection tests for SARS-CoV-2 including any of the codes below:

94307-6: SARS-CoV-2 (COVID-19) N gene [Presence] in Specimen by Nucleic acid amplification using CDC primer-probe set N110

94308-4: SARS-CoV-2 (COVID-19) N gene [Presence] in Specimen by Nucleic acid amplification using CDC primer-probe set N213

94309-2: SARS-CoV-2 (COVID-19) RNA [Presence] in Specimen by NAA with probe detection2,104,487

94310-0: SARS-related coronavirus N gene [Presence] in Specimen by Nucleic acid amplification using CDC primer-probe set N311

94314-2: SARS-CoV-2 (COVID-19) RdRp gene [Presence] in Specimen by NAA with probe detection12,231

94315-9: SARS-related coronavirus E gene [Presence] in Specimen by NAA with probe detection10

94316-7: SARS-CoV-2 (COVID-19) N gene [Presence] in Specimen by NAA with probe detection118,417

94500-6: SARS-CoV-2 (COVID-19) RNA [Presence] in Respiratory specimen by NAA with probe detection4,746,972

94502-2: SARS-related coronavirus RNA [Presence] in Respiratory specimen by NAA with probe detection11,974

94533-7: SARS-CoV-2 (COVID-19) N gene [Presence] in Respiratory specimen by NAA with probe detection51,554

94534-5: SARS-CoV-2 (COVID-19) RdRp gene [Presence] in Respiratory specimen by NAA with probe detection332,029

94559-2: SARS-CoV-2 (COVID-19) ORF1ab region [Presence] in Respiratory specimen by NAA with probe detection48,000

94565-9: SARS-CoV-2 (COVID-19) RNA [Presence] in Nasopharynx by NAA with non-probe detection84,044

94759-8: SARS-CoV-2 (COVID-19) RNA [Presence] in Nasopharynx by NAA with probe detection42,222

94760-6: SARS-CoV-2 (COVID-19) N gene [Presence] in Nasopharynx by NAA with probe detection13,728

94845-5: SARS-CoV-2 (COVID-19) RNA [Presence] in Saliva (oral fluid) by NAA with probe detection28,110

95406-5: SARS-CoV-2 (COVID-19) RNA [Presence] in Nose by NAA with probe detection8,078

95409-9: SARS-CoV-2 (COVID-19) N gene [Presence] in Nose by NAA with probe detection127

96123-5: SARS-CoV-2 (COVID-19) RdRp gene [Presence] in Upper respiratory specimen by NAA with probe detection

LOINC 94558-4: SARS-CoV-2 (COVID-19) Ag [Presence] in Respiratory specimen by Rapid immunoassay

LOINC 94763-0: SARS-CoV-2 (COVID-19) Ag [Presence] in Specimen by Organism specific culture

LOINC 94558-4: SARS-CoV-2+SARS-CoV-2 (COVID-19) Ag [Presence] in Respiratory specimen by Rapid immunoassay

LOINC 96119-3: SARS-CoV-2 (COVID-19) Ag [Presence] in Upper respiratory specimen by Immunoassay

LOINC 96119-3: SARS-CoV-2 (COVID-19) Ag [Presence] in Upper respiratory specimen by Rapid immunoassay

TNX curated 9047 is a composite code for Aspartate aminotransferase {Enzymatic activity/volume] in Serum or Plasma includes any of the codes below:

LOINC 1920-8: Aspartate aminotransferase [Enzymatic activity/volume] in Serum or Plasma

LOINC 30239-8: Aspartate aminotransferase [Enzymatic activity/volume] in Serum or Plasma by With P-5'-P

TNX curated 9044 is a composite code for Alanine aminotransferase [Enzymatic activity/volume] in Serum, Plasma or Blood includes any of the codes below:

LOINC 1742-6: Alanine aminotransferase [Enzymatic activity/volume] in Serum or Plasma

LOINC 1743-4: Alanine aminotransferase [Enzymatic activity/volume] in Serum or Plasma by With P-5'-P

LOINC 1744-2: Alanine aminotransferase [Enzymatic activity/volume] in Serum or Plasma by No addition of P-5'-P

LOINC 76625-3: Alanine aminotransferase [Enzymatic activity/volume] in Blood

LOINC 77144-4: Alanine aminotransferase [Enzymatic activity/volume] in Serum, Plasma or Blood

TNX curated 9050 is a composite code for Bilirubin total [Mass/volume] in Serum, Plasma or Blood includes any of the codes below:

LOINC 1975-2: Bilirubin.total [Mass/volume] in Serum or Plasma

LOINC 42719-5: Bilirubin.total [Mass/volume] in Blood

TNX curated 9089 is a composite code for SARS coronavirus 2 IgG IgM Ab [Presence] in Serum or Plasma includes any of the codes below:

LOINC 94507-1: SARS-CoV-2 (COVID-19) IgG Ab [Presence] in Serum, Plasma or Blood by Rapid immunoassay

LOINC 94508-9: SARS-CoV-2 (COVID-19) IgM Ab [Presence] in Serum, Plasma or Blood by Rapid immunoassay

LOINC 94547-7: SARS-CoV-2 (COVID-19) IgG+IgM Ab [Presence] in Serum or Plasma by Immunoassay

LOINC 94563-4: SARS-CoV-2 (COVID-19) IgG Ab [Presence] in Serum or Plasma by Immunoassay

LOINC 94564-2: SARS-CoV-2 (COVID-19) IgM Ab [Presence] in Serum or Plasma by Immunoassay

LOINC 94761-4: SARS-CoV-2 (COVID-19) IgG Ab [Presence] in DBS by Immunoassay

LOINC 99596-9: SARS-CoV-2 (COVID-19) N protein IgG Ab [Presence] in Serum or Plasma by Immunoassay

LOINC 99597-7: SARS-CoV-2 (COVID-19) S protein IgG Ab [Presence] in Serum or Plasma by Immunoassay

Matching codes:

Age: Current age

AI: Age at index

M: Male

F: Female

UN: Unknown gender

2186-5: Not Hispanic or Latino

2134-2: Hispanic or Latino

UN: Unknown ethnicity

2106-3: White

2054-5: Black or African American

2028-9: Asian

1002-5: Native American/Alaska native

2076-8: Native Hawaiian or Other Pacific Islander

2131-1: Unknown race

Z23: Encounter for immunization

Z68.5: Body mass index [BMI] pediatric

Z68.51: Body mass index [BMI] pediatric, less than 5th percentile for age

Z68.52: Body mass index [BMI] pediatric, 5th percentile to less than 85th percentile for age

Z68.53: Body mass index [BMI] pediatric, 85th percentile to less than 95th percentile for age

Z68.54: Body mass index [BMI] pediatric, greater than or equal to 95th percentile for age

1013829: Preventive medicine services

91300: Severe acute respiratory syndrome coronavirus 2 (SARS-CoV-2) (Coronavirus disease [COVID-19]) vaccine, mRNA-LNP, spike protein, preservative free, 30 mcg/0.3mL dosage, diluent reconstituted, for intramuscular use

| **eTable 1. Baseline Characteristics of Study Population Before and After Matching (Age 1-4 Years)** | | | | | | |
| --- | --- | --- | --- | --- | --- | --- |
|  | Unmatched cohort, No. (%) | | | Matched cohort, No. (%) | | |
|  | COVID-19  n=119,455 | ORI  n=601,209 | SMD | COVID-19  n=119,379 | ORI  n=119,379 | SMD |
| Age at Index in years, mean (SD) | 2.33±1.20 | 2.18±1.14 | 0.12 | 2.32±1.19 | 2.31±1.14 | 0.003 |
| Sex |  |  |  |  |  |  |
| Male | 63,621 (53.3) | 321,076 (53.4) | <0.01 | 63,588 (53.3) | 63,557 (53.2) | <0.001 |
| Female | 55,763 (46.7) | 279,941 (46.6) | <0.01 | 55,727 (46.7) | 55,758 (46.7) | <0.001 |
| Unknown | 71 (0.1) | 192 (<0.1) | 0.01 | 64 (0.1) | 64 (0.1) | <0.001 |
| Ethnicity |  |  |  |  |  |  |
| Not Hispanic | 73,625 (61.6) | 370,871 (61.7) | <0.01 | 73,576 (61.6) | 73,550 (61.6) | <0.001 |
| Hispanic | 20,717 (17.3) | 104,616 (17.4) | <0.01 | 20,696 (17.3) | 20,719 (17.3) | <0.001 |
| Unknown | 25,113 (21.0) | 125,722 (20.9) | <0.01 | 25,107 (21.0) | 25,110 (21.0) | <0.001 |
| Race |  |  |  |  |  |  |
| White | 61,653 (51.6) | 314,847 (52.4) | 0.02 | 61,610 (51.6) | 61,579 (51.6) | <0.001 |
| Black | 23.707 (19.8) | 116,257 (19.3) | 0.01 | 23,688 (19.8) | 23,674 (19.8) | <0.001 |
| Asian | 5052 (4.2) | 23,633 (3.9) | 0.01 | 5050 (4.2) | 5091 (3.3) | 0.002 |
| Native American or Alaska Native | 597 (0.5) | 3362 (0.6) | 0.01 | 597 (0.5) | 587 (0.5) | 0.001 |
| Native Hawaiian or Other Pacific Islander | 512 (0.4) | 2900 (0.5) | 0.01 | 512 (0.4) | 513 (0.4) | <0.001 |
| Unknown Race | 27,934 (23.4) | 140,210 (23.3) | <0.01 | 27,922 (23.4) | 27,935 (23.4) | <0.001 |
| BMI, pediatric |  |  |  |  |  |  |
| measured | 10,503 (8.8) | 230,511 (38.3) | 0.01 | 10,480 (8.8) | 10,452 (8.8) | 0.001 |
| <5^th^ percentile | 998 (0.8) | 4712 (0.8) | 0.02 | 995 (0.8z0 | 934 (0.8) | 0.006 |
| 5^th^ to <85^th^ percentile | 7566 (6.3) | 41,029 (6.8) | <0.01 | 7561 (6.3) | 7537 (6.3) | 0.001 |
| 85^th^ to <95^th^ percentile | 1723 (1.4) | 8441 (1.4) | <0.01 | 1718 (1.4) | 1695 (1.4) | 0.002 |
| ≥95^th^ percentile | 1969 (1.6) | 8949 (1.5) | 0.01 | 1954 (1.6) | 1961 (1.6) | 0.001 |
| Preventive medicine services | 44,338 (37.1) | 239,056 (39.8) | 0.05 | 44,323 (37.1) | 44,460 (37.2) | 0.002 |
| Encounter for immunization | 41,480 (34.7) | 230,511 (38.3) | 0.08 | 41,463 (34.7) | 41,516 (34.8) | 0.001 |
| SARS-CoV-2 vaccination | 235 (0.2) | 1580 (0.3) | 0.01 | 235 (0.2) | 201 (0.2) | 0.007 |
| Abbreviations: ORI, other respiratory infection; SMD, standard mean difference; SD, standard deviation | | | | | | |

| **eTable 2. Baseline Characteristics of Study Population Before and After Matching with Inpatient Stay (Age 1-10 Years)** | | | | | | |
| --- | --- | --- | --- | --- | --- | --- |
|  | Unmatched cohort, No. (%) | | | Matched cohort, No. (%) | | |
|  | COVID-19  n=10,257 | ORI  n=44,970 | SMD | COVID-19  n=10,256 | ORI  n=10,256 | SMD |
| Age at Index in years, mean (SD) | 4.59±3.06 | 3.56±2.69 | 0.36 | 4.59±3.06 | 4.55±3.04 | 0.013 |
| Sex |  |  |  |  |  |  |
| Male | 5628 (54.9) | 24,930 (55.4) | 0.01 | 5628 (54.9) | 5608 (54.7) | 0.004 |
| Female | 4626 (45.1) | 20,034 (44.6) | 0.01 | 4625 (45.1) | 4647 (45.3) | 0.004 |
| Unknown | 10 | 10 |  | 10 | 10 |  |
| Ethnicity |  |  |  |  |  |  |
| Not Hispanic | 6612 (64.5) | 30,457 (67.7) | 0.07 | 6612 (64.5) | 6595 (64.3) | 0.004 |
| Hispanic | 2095 (20.4) | 7346 (16.3) | 0.11 | 2094 (20.4) | 2119 (20.7) | 0.006 |
| Unknown | 1550 (15.1) | 7167 (15.9) | 0.02 | 1550 (15.1) | 1542 (15.0) | 0.002 |
| Race |  |  |  |  |  |  |
| White | 5164 (50.3) | 24,252 (53.9) | 0.07 | 5164 (50.4) | 5212 (50.8) | 0.009 |
| Black | 2151 (21.0) | 9078 (20.2) | 0.02 | 2150 (21.0) | 2161 (21.1) | 0.003 |
| Asian | 330 (3.2) | 1407 (3.1) | 0.01 | 330 (3.2) | 311 (3.0) | 0.011 |
| Native American or Alaska Native | 92 (0.9) | 494 (1.1) | 0.02 | 92 (0.9) | 68 (0.7) | 0.027 |
| Native Hawaiian or Other Pacific Islander | 25 (0.2) | 119 (0.3) | <0.01 | 25 (0.2) | 24 (0.2) | 0.002 |
| Unknown Race | 2495 (24.3) | 9620 (21.4) | 0.07 | 2495 (24.3) | 2480 (24.2) | 0.003 |
| BMI, pediatric |  |  |  |  |  |  |
| measured | 1352 (13.2) | 8119 (18.1) | 0.13 | 1352 (13.2) | 1310 (12.8) | 0.012 |
| <5^th^ percentile | 184 (1.8) | 800 (1.8) | <0.01 | 184 (1.8) | 150 (1.5) | 0.026 |
| 5^th^ to <85^th^ percentile | 860 (8.4) | 5621 (12.5) | 0.13 | 860 (8.4) | 835 (8.1) | 0.009 |
| 85^th^ to <95^th^ percentile | 326 (3.2) | 1841 (4.1) | 0.05 | 326 (3.2) | 306 (3.0) | 0.011 |
| ≥95^th^ percentile | 428 (4.2) | 2067 (4.6) | 0.05 | 326 (3.2) | 306 (3.0) | 0.011 |
| Preventive medicine services | 3247 (31.7) | 16,022 (35.6) | 0.08 | 3246 (31.6) | 3284 (32.0) | 0.008 |
| Encounter for immunization | 2710 (26.4) | 14,572 (32.4) | 0.13 | 2710 (26.4) | 2704 (26.4) | 0.001 |
| SARS-CoV-2 vaccination | 52 (0.5) | 290 (0.6) | 0.02 | 52 (0.5) | 35 (0.3) | 0.026 |
| Abbreviations: ORI, other respiratory infection; SMD, standard mean difference; SD, standard deviation | | | | | | |

| **eTable 3. Baseline Characteristics of Study Population Before and After Matching with Inpatient Stay (Age 1-4 Years)** | | | | | | |
| --- | --- | --- | --- | --- | --- | --- |
|  | Unmatched cohort, No. (%) | | | Matched cohort, No. (%) | | |
|  | COVID-19  n=5575 | ORI  n=30,807 | SMD | COVID-19  n=5574 | ORI  n=5574 | SMD |
| Age at Index in years, mean (SD) | 2.12±1.13 | 1.96±1.07 | 0.14 | 2.12±1.13 | 2.12±1.12 | 0.003 |
| Sex |  |  |  |  |  |  |
| Male | 3099 (55.6) | 17,438 (56.6) | 0.02 | 3098 (55.6) | 3085 (55.3) | 0.004 |
| Female | 2475 (44.4) | 13,365 (43.4) | 0.02 | 2475 (44.4) | 2489 (44.7) | 0.005 |
| Unknown | 10 | 10 |  | 10 | 0 |  |
| Ethnicity |  |  |  |  |  |  |
| Not Hispanic | 3512 (63.0) | 20,511 (66.6) | 0.08 | 3512 (63.0) | 3498 (62.8) | 0.005 |
| Hispanic | 1133 (20.3) | 4962 (16.1) | 0.11 | 1132 (20.3) | 1146 (20.6) | 0.006 |
| Unknown | 930 (16.7) | 5334 (17.3) | 0.02 | 930 (16.7) | 930 (16.7) | <0.001 |
| Race |  |  |  |  |  |  |
| White | 2709 (48.6) | 16,096 (52.2) | 0.07 | 2709 (48.6) | 2736 (49.1) | 0.010 |
| Black | 1133 (20.0) | 6129 (19.9) | <0.01 | 1112 (20.0) | 1106 (19.8) | 0.003 |
| Asian | 201 (3.6) | 1030 (3.3) | 0.01 | 201 (3.6) | 209 (3.8) | 0.008 |
| Native American or Alaska Native | 47 (0.8) | 363 (1.2) | 0.03 | 47 (0.2) | 42 (0.8) | 0.010 |
| Native Hawaiian or Other Pacific Islander | 13 (0.2) | 83 (0.3) | 0.01 | 13 | 10 |  |
| Unknown Race | 1492 (26.8) | 7106 (23.1) | 0.09 | 1492 (26.8) | 1476 (26.5) | 0.006 |
| BMI, pediatric |  |  |  |  |  |  |
| measured | 412 (7.4) | 3143 (10.2) | 0.10 | 412 (7.4) | 375 (6.7) | 0.026 |
| <5^th^ percentile | 75 (1.3) | 382 (1.2) | 0.01 | 75 (1.3) | 53 (1.0) | 0.037 |
| 5^th^ to <85^th^ percentile | 272 (4.9) | 2237 (7.3) | 0.10 | 272 (4.9) | 260 (4.7) | 0.010 |
| 85^th^ to <95^th^ percentile | 76 (1.4) | 537 (1.7) | 0.03 | 76 (1.4) | 79 (1.4) | 0.005 |
| ≥95^th^ percentile | 78 (1.4) | 561 (1.8) | 0.03 | 78 (1.4) | 66 (1.2) | 0.019 |
| Preventive medicine services | 1544 (27.7) | 9491 (30.8) | 0.07 | 1543 (27.7) | 1559 (28.0) | 0.006 |
| Encounter for immunization | 1361 (24.4) | 8901 (28.9) | 0.08 | 1361 (24.4) | 1367 (24.5) | 0.002 |
| SARS-CoV-2 vaccination | 10 | 96 |  | 10 | 10 |  |
| Abbreviations: ORI, other respiratory infection; SMD, standard mean difference; SD, standard deviation | | | | | | |

| **eTable 4.** **Comparison of clinical outcomes during 1 to 6 months after COVID-19 vs ORI among matched cohorts of patients aged 1-10 years (01/01/2020 – 12/31/2022), not excluding those with liver abnormalities in the acute phase** | | | | | |
| --- | --- | --- | --- | --- | --- |
| n, All 1-4 = 119,379  n, All 1-10 = 260,132  n, Inpt 1-4 = 5575  n, Inpt 1-10 = 10,256 |  | Age Group  (years) | COVID-19 Cohort No. | ORI Cohort No. | RR (95%CI) |
| **Elevated AST**^a^ **or ALT**^b^ | All | 1-4 | 245 | 82 | 2.99 (2.33-3.84) |
|  |  | 1-10 | 472 | 144 | 3.28 (2.72-3.95) |
|  | Inpt | 1-4 | 156 | 44 | 3.45 (2.54-4.94) |
|  |  | 1-10 | 274 | 70 | 3.91 (3.02-5.08) |
| **Elevated TBil**^c^ | All | 1-4 | 74 | 21 | 3.52 (2.17-5.72) |
|  |  | 1-10 | 148 | 31 | 4.77 (3.24-7.03) |
|  | Inpt | 1-4 | 41 | 12 | 3.42 (1.80-6.49) |
|  |  | 1-10 | 72 | 16 | 4.50 (2.62-7.73) |
| **Diseases of the liver (K70-K77) OR viral hepatitis (B15-B19)** | All | 1-4 | 71 | 29 | 2.45 (1.59-3.77) |
|  |  | 1-10 | 170 | 85 | 2.00 (1.54-2.60) |
|  | Inpt | 1-4 | 40 | 13 | 3.08 (1.65-5.75) |
|  |  | 1-10 | 72 | 15 | 4.80 (2.75-8.37) |
| **Diseases of the liver (K70-K77)** | All | 1-4 | 60 | 17 | 3.53 (2.06-6.05) |
|  |  | 1-10 | 156 | 69 | 2.26 (1.70-3.00) |
|  | Inpt | 1-4 | 39 | 13 | 3.00 (1.60-5.61) |
|  |  | 1-10 | 69 | 14 | 4.93 (2.78-8.75) |
| Other inflammatory disease of the liver (K75) | All | 1-4 | 16 | 10 | 1.60 (0.73-3.53) |
|  |  | 1-10 | 31 | 11 | 2.82 (1.42-5.61) |
|  | Inpt | 1-4 | 14 | 10 | 1.40 (0.62-3.15) |
|  |  | 1-10 | 23 | 10 | 2.30 (1.10-4.83) |
| Other diseases of the liver (K76) | All | 1-4 | 40 | 13 | 3.08 (1.65-5.75) |
|  |  | 1-10 | 120 | 60 | 2.00 (1.47-2.73) |
|  | Inpt | 1-4 | 24 | 10 | 2.40 (1.15-5.01) |
|  |  | 1-10 | 40 | 10 | 4.60 (2.32-9.11) |
| **Hepatitis of unspecified etiology**^d^ | All | 1-4 | 20 | 10 | 2.00 (0.94-4.27) |
|  |  | 1-10 | 32 | 13 | 2.83 (1.29-4.69) |
|  | Inpt | 1-4 | 17 | 10 | 1.70 (0.78-3.71) |
|  |  | 1-10 | 22 | 10 | 2.20 (1.04-4.64) |

Abbreviations: ORI, other respiratory infection; RR, risk ratio; CI, confidence interval; AST, aspartate aminotransferase; ALT, alanine aminotransferase; TBil, total bilirubin; Inpt, inpatient. Cohorts matched on factors listed in Table 1. Patients with pre-existing disease are excluded from the cohorts; however, patients with the outcome during the acute period (from 1 months before to month after COVID-19 or ORI indexed encounter diagnosis) are NOT excluded from the analysis.

^a^ AST ≥110 units per liter

^b^ ALT ≥100 units per liter

^c^ TBil ≥2 milligrams per deciliter

^d^ Surveillance codes include B17.8, B17.9, B19.0, B19.9, K71.6, K72.0, K72.9, K75.2, K75.9

| **eTable 5.** **Comparison of clinical outcomes during 1 to 6 months after COVID-19 vs ORI among matched cohorts of patients aged 1-10 years (01/01/2020 – 12/31/2022), not excluding those with previous liver abnormalities** | | | | | |
| --- | --- | --- | --- | --- | --- |
| n, All 1-4 = 129,111  n, All 1-10 = 279,161  n, Inpt 1-4 = 6520  n, Inpt 1-10 = 11,995 |  | Age Group  (years) | COVID-19 Cohort No. | ORI Cohort No. | RR (95%CI) |
| **Elevated AST**^a^ **or ALT**^b^ | All | 1-4 | 468 | 127 | 3.68 (3.03-4.48) |
|  |  | 1-10 | 930 | 267 | 3.48 (3.04-3.99) |
|  | Inpt | 1-4 | 320 | 77 | 4.16 (3.25-5.32) |
|  |  | 1-10 | 589 | 171 | 3.44 (2.91-4.08) |
| **Elevated TBil**^c^ | All | 1-4 | 161 | 45 | 3.58 (2.57-4.98) |
|  |  | 1-10 | 383 | 117 | 3.27 (2.66-4.03) |
|  | Inpt | 1-4 | 92 | 22 | 4.18 (2.63-6.65) |
|  |  | 1-10 | 204 | 74 | 2.76 (2.12-3.59) |
| **Diseases of the liver (K70-K77) OR viral hepatitis (B15-B19)** | All | 1-4 | 159 | 55 | 2.89 (2.13-3.93) |
|  |  | 1-10 | 367 | 165 | 2.22 (1.85-2.67) |
|  | Inpt | 1-4 | 95 | 22 | 4.32 (2.72-6.86) |
|  |  | 1-10 | 158 | 49 | 3.22 (2.34-4.44) |
| **Viral hepatitis (B15-B19)** | All | 1-4 | 32 | 13 | 2.46 (1.29-4.69) |
|  |  | 1-10 | 42 | 17 | 2.47 (1.41-4.34) |
|  | Inpt | 1-4 | * | * | * |
|  |  | 1-10 | * | * | * |
| **Diseases of the liver (K70-K77)** | All | 1-4 | 132 | 43 | 3.07 (2.18-4.33) |
|  |  | 1-10 | 333 | 151 | 2.21 (1.82-2.67) |
|  | Inpt | 1-4 | 92 | 22 | 4.18 (2.63-6.65) |
|  |  | 1-10 | 153 | 49 | 3.12 (2.27-4.30) |
| Hepatic failure, not elsewhere classified (K72) | All | 1-4 | 24 | 10 | 2.40 (1.15-5.02) |
|  |  | 1-10 | 41 | 21 | 1.95 (1.15-3.30) |
|  | Inpt | 1-4 | 18 | 11 | 1.64 (0.77-3.46) |
|  |  | 1-10 | 22 | 12 | 1.83 (0.91-3.70) |
| Other inflammatory disease of the liver (K75) | All | 1-4 | 30 | 10 | 3.00 (1.47-6.14) |
|  |  | 1-10 | 70 | 16 | 4.38 (2.54-7.53) |
|  | Inpt | 1-4 | 23 | 10 | 2.30 (1.10-4.83) |
|  |  | 1-10 | 41 | 11 | 3.73 (1.92-7.25) |
| Other diseases of the liver (K76) | All | 1-4 | 89 | 32 | 2.78 (1.86-4.16) |
|  |  | 1-10 | 242 | 124 | 1.95 (1.57-2.42) |
|  | Inpt | 1-4 | 61 | 10 | 6.10 (3.13-11.90) |
|  |  | 1-10 | 103 | 34 | 3.03 (2.06-4.46) |
| **Hepatitis of unspecified etiology**^d^ | All | 1-4 | 44 | 10 | 4.40 (2.21-8.74) |
|  |  | 1-10 | 66 | 27 | 2.44 (1.56-3.83) |
|  | Inpt | 1-4 | 34 | 11 | 3.09 (1.57-6.10) |
|  |  | 1-10 | 44 | 15 | 2.93 (1.63-5.27) |

Abbreviations: ORI, other respiratory infection; RR, risk ratio; CI, confidence interval; AST, aspartate aminotransferase; ALT, alanine aminotransferase; TBil, total bilirubin; Inpt, inpatient. Cohorts matched on factors listed in Table 1. Patients with pre-existing disease are NOT excluded from the cohorts; patients with the outcome during the acute period (from 1 months before to month after COVID-19 or ORI indexed encounter diagnosis) are also NOT excluded from the analysis.

^a^ AST ≥110 units per liter

^b^ ALT ≥100 units per liter

^c^ TBil ≥2 milligrams per deciliter

^d^ Surveillance codes include B17.8, B17.9, B19.0, B19.9, K71.6, K72.0, K72.9, K75.2, K75.9

*Indicates counts too few to report

**eTable 6.** **Comparison of elevated laboratory values during 1 to 6 months after COVID-19 vs ORI among matched cohorts of pediatric patients with laboratory values measured during the acute period of infection (01/01/2020 – 12/31/2022)**

| n, (1-4) = 5513  n, (1-10) = 11,210 | Age Group | COVID-19 Cohort  No. | ORI Cohort  No. | RR (95% CI) |
| --- | --- | --- | --- | --- |
| **Elevated AST**^a^ **or ALT**^b^ | 1-4 | 82 | 37 | 2.23 (1.50-3.27) |
|  | 1-10 | 334 | 107 | 3.12 (2.51-3.88) |
| **Elevated TBil**^c^ | 1-4 | 19 | 10 | 3.61 (1.35-9.66) |
|  | 1-10 | 97 | 38 | 2.55 (1.76-3.71) |

Abbreviations: ORI, other respiratory infection; HR, hazard ratio; CI, confidence interval; AST, aspartate aminotransferase; ALT, alanine aminotransferase; TBil, total bilirubin; Inpt, inpatient. Cohorts matched on factors listed in Table 1. Patients with elevated labs from 0 to 1 months after respiratory illness are NOT excluded from this analysis.

^a^ AST ≥110 units per liter

^b^ ALT ≥100 units per liter

^c^ TBil ≥2 milligrams per deciliter

| **eTable 7. Comparison of pre-existing conditions among cases with hepatic complications**^a^ **after COVID-10 vs ORI aged 1 – 10 years (01/01/2020 – 12/31/2022)** | | | | | | |
| --- | --- | --- | --- | --- | --- | --- |
|  | All Cases | | Cases with Inpatient Stay | | | |
|  | COVID-19  (%) | ORI  (%) | SMD | COVID-19  (%) | ORI  (%) | SMD |
| Age at Index in years, mean (SD) | 5.51±3.24 | 4.42±3.21 | 0.34 | 4.70±3.02 | 4.29±3.20 | 0.13 |
| Sex |  |  |  |  |  |  |
| Male | 61 | 58 | 0.06 | 57 | 56 | 0.02 |
| Female | 39 | 42 | 0.06 | 43 | 44 | 0.02 |
| Unknown | N/A | N/A | N/A | N/A | N/A | N/A |
| Ethnicity |  |  |  |  |  |  |
| Not Hispanic | 63 | 59 | 0.07 | 63 | 75 | 0.26 |
| Hispanic | 27 | 25 | 0.05 | 19 | 11 | 0.22 |
| Unknown | 14 | 26 | 0.16 | 18 | 14 | 0.11 |
| Race |  |  |  |  |  |  |
| White | 49 | 60 | 0.22 | 42 | 56 | 0.27 |
| Black | 21 | 12 | 0.24 | 27 | 17 | 0.24 |
| Asian | * | * | * | * | * | * |
| Native American or Alaska Native | * | * | * | * | * | * |
| Native Hawaiian or Other Pacific Islander | * | * | * | * | * | * |
| Unknown Race | 24 | 23 | 0.02 | 27 | 21 | 0.13 |
| BMI, pediatric |  |  |  |  |  |  |
| measured | 31 | 11 | 0.50 | 16 | 26 | 0.26 |
| <5^th^ percentile | 1 | 1 | <0.01 | * | * | * |
| 5^th^ to <85^th^ percentile | 10 | 6 | 0.14 | * | * | * |
| 85^th^ to <95^th^ percentile | * | * | * | * | * | * |
| ≥95^th^ percentile | 20 | 7 | 0.38 | * | * | * |
| Preventive medicine services | 31 | 20 | 0.26 | 22 | 27 | 0.12 |
| Encounter for immunization | 34 | 19 | 0.36 | 24 | 35 | 0.24 |
| SARS-CoV-2 vaccination | * | * | * | * | * | * |
| A00-B99 Certain infectious and parasitic diseases | 46 | 27 | 0.40 | 46 | 50 | 0.08 |
| C00-D49 Neoplasm | 15 | 8 | 0.22 | 20 | 12 | 0.22 |
| D50-D89 Diseases of the blood and blood-forming organs and certain disorders involving the immune mechanism | 34 | 13 | 0.49 | 43 | 30 | 0.29 |
| E00-E89 Endocrine, nutritional and metabolic diseases | 68 | 40 | 0.57 | 54 | 47 | 0.15 |
| F01-F99 Mental, Behavioral and Neurodevelopmental disorders | 31 | 18 | 0.31 | 33 | 37 | 0.08 |
| G00-G99 Diseases of the nervous system | 33 | 16 | 0.39 | 29 | 41 | 0.25 |
| H00-H59 Diseases of the eye and adnexa | 34 | 14 | 0.49 | 30 | 38 | 0.17 |
| H60-H95 Diseases of the ear and mastoid process | 43 | 21 | 0.47 | 36 | 37 | 0.01 |
| I00-I99 Diseases of the circulatory system | 24 | 15 | 0.22 | 30 | 38 | 0.17 |
| J00-J99 Diseases of the respiratory system | 59 | 31 | 0.59 | 52 | 70 | 0.39 |
| K00-K95 Diseases of the digestive system | 51 | 36 | 0.32 | 52 | 62 | 0.20 |
| L00-L99 Diseases of the skin and subcutaneous tissue | 43 | 24 | 0.40 | 42 | 43 | 0.01 |
| M00-M99 Diseases of the musculoskeletal system and connective tissue | 30 | 15 | 0.36 | 29 | 33 | 0.09 |
| N00-N99 Diseases of the genitourinary system | 24 | 15 | 0.24 | 24 | 26 | 0.05 |
| P00-P96 Certain conditions originating in the perinatal period | 13 | 16 | 0.08 | 16 | 30 | 0.36 |
| Q00-Q99 Congenital malformations, deformations and chromosomal abnormalities | 27 | 27 | <0.01 | 31 | 48 | 0.34 |
| S00-T88 Injury, poisoning and certain other consequences of external causes | 40 | 27 | 0.28 | 43 | 46 | 0.05 |
| V00-Y99 External causes of morbidity | 18 | 8 | 0.28 | 18 | 19 | 0.03 |
| Abbreviations: ORI, other respiratory infection; SMD, standard mean difference; SD, standard deviation  Shading highlights the cohort with the higher proportion when SMD is ≥ 0.1  ^a^ Any encounter diagnosis of K70-K77 or B15-B19 during the 1-6 month follow-up period after respiratory infection  *Indicates counts too small to calculate % | | | | | | |
